# Global Burden of Disease worldwide statistical analysis of risk factors impacting noncommunicable disease deaths compared with assessments by systematic literature reviews

**DOI:** 10.1101/2020.12.02.20242867

**Authors:** David K Cundiff, Chunyi Wu

## Abstract

**Background:** Based on systematic literature reviews, the Institute of Health Metrics and Evaluation (IHME) Global Burden of Disease (GBD) landmark risk factor paper attributes about 8 million noncommunicable diseases deaths/year to a composite of 15 dietary risk factors. Another 27 million noncommunicable deaths out of 56.5 million total deaths worldwide are attributed to high levels of metabolic risk factors.

**Methods:** We format and population weight IHME GBD worldwide data resulting in7886 rows of data from cohorts of about 1 million people each representing about 7.8 billion people in 2020. Noncommunicable disease deaths/100k/year (NCD) are correlated with the four metabolic risk factors, 15 dietary risk factors, six dietary covariates, and the composite “dietary risks” variable. We compare NCD groups of cohorts (about 1000 cohorts, representing about 1 billion people) with the lowest and highest NCD and the same for body mass index (BMI). We compare literature reviews attributions of NCD causation with findings from the GBD data in assessing the impact of dietary and metabolic risk factors on NCD.

**Findings:** Literature review derived dietary risks composite risk factor correlated with NCD account for 1.9% of worldwide NCD. Using GBD data to compare groups with the lowest versus the highest rates of NCD: the lowest NCD 990 cohorts average 923.8 NCD and consume 320.4 kcal/day from six animal foods. The highest NCD 1054 cohorts average 2064 NCD and on average and consume 117.2 kcal/day of six animal foods. Vitamin A deficiency in children is 75% higher in children in the high NCD cohorts. Evaluating high BMI cohorts, 973 cohorts with BMI≥25 account for 10.5% of the worldwide NCD. These high BMI cohorts also have higher than average fasting plasma glucose (FPG: FPG mean=4.71 versus FPG mean=4.25) and low density lipoprotein cholesterol (LDLC: LDLC mean=2.79 versus LDLC mean=2.30).

**Interpretation:** The evidence basis of the dietary risks composite risk factor, accounting for only 1.9% of NCD worldwide should be questioned. Systematic literature reviews based versus GBD data based estimates of metabolic and dietary risk factor impacts on NCD vary markedly. Systematic literature reviews of studies involving individual subjects are better at ascertaining the impact on NCD of obesity, hyperglycemia, high LDLC, and hypertension. High levels of metabolic risk factors cannot be analysed with GBD data giving only the mean values for NCD and risk factors. Since high mean BMI cohorts (mean BMI≥25) account for only 10.5% of worldwide NCD, literature reviews appear to overestimate the worldwide impacts of high metabolic risk factors on NCD (i.e., accounting for nearly 50% of worldwide deaths: 27 million/56.5 million worldwide deaths). High NCD, including cardiovascular disease deaths, associated with low animal food intake suggests that fat soluble vitamin deficiencies (vitamins A, D, E and K2, all largely from animal foods that also supply fatty acids for gut absorption) may partly account for high rates of NCD worldwide, especially in developing countries. Literature review based and GBD raw data based approaches should be complementary in assessing dietary and other attributable risks for NCD and other individual health outcomes.

**Funding:** None

**Research in context:** *Evidence before this study:* The Global Burden of Diseases, Injuries, and Risk Factors Study (GBD 2019) provides the most recent assessment of deaths, attributable to metabolic, environmental and occupational, and behavioural risk factors. Controversies abound in the field of nutritional epidemiology. The lipid hypothesis that dietary saturated fat and cholesterol promote cardiovascular diseases has been disputed with no scientific consensus on the outcome.

*Added value of this study:* This study assesses the evidence basis of deaths attributed to dietary risks and metabolic risk factors derived from systematic reviews of the medical literature in GBD 2019. This study uses GBD raw data formatted and population weighted to assess the worldwide deaths from noncommunicable diseases attributable to dietary and metabolic risk factors. With this GBD data analysis methodology, the levels of 20 dietary risk factors are assessed in cohorts with the highest and lowest rates of noncommunicable disease deaths. Worldwide GBD data show that many deaths are associated with low levels of animal foods and saturated fatty acids (kcal/day percapita) and associated with low levels of metabolic risk factors. These findings are at variance with the lipid hypothesis operating uniformly worldwide. In the UK, USA, Mexico, and Japan, with subnational GBD data, many deaths are attributable to high levels of animal foods, saturated fatty acids, and metabolic risk factors, supporting the lipid hypothesis.

*Implications of all the available evidence:* The findings of systematic literature reviews in assessing the health impacts of high metabolic risk factors are superior than using GBD data analysis because GBD data only assess mean cohort levels of metabolic risk factors. However, GBD data can show effects of dietary risk factors acting directly and mediating metabolic risks leading to noncommunicable disease deaths. This GBD data based methodology can enhance understanding of the complex interrelationships of diet, metabolic risk factors and noncommunicable disease deaths. The findings are consistent with a “fat soluble vitamin hypothesis” that deficiencies of vitamins A, D, E, and K2 and fatty acids from inadequate animal food consumption lead to increased cardiovascular disease and noncommunicable disease deaths.

## Introduction

Institute of Health Metrics and Evaluation (IHME) Global Burden of Disease (GBD) researchers attribute about 8 million deaths/year worldwide to poor diet quality out of a total of about 56.5 million deaths a year worldwide.^1^ GBD investigators ascribe high body mass index (BMI≥25) to poor “diet quality,” excessive caloric intake, and low physical activity. Diet quality is defined as, “the dietary risks that are based on the joint effects of 15 diet quality components.” Each dietary risk factor has an optimal range or theoretical minimum risk exposure level range (TMREL range in g/day) to consume based on systematic reviews of the literature. “Dietary risks” is a composite metric including harmful risk factors: (1) high processed meat (TMREL=0), (2) high red meat (TMREL=0), (3) high sugary beverages (TMREL=Any intake in g/day of beverages with ≥50 kcal per 226.8 gram serving), (4) high trans fatty acids (TMREL=0), and (5) high sodium (TMREL=average 24-hour urinary sodium excretion of 1-5 g/day) and protective risk factors: (6) low milk (TMREL=90-100 g/day), (7) low omega-3 fatty acids (TMREL= 430-470 milligrams of eicosapentaenoic acid (EPA) and docosahexaenoic acid (DHA)), (8) low fruits (TMREL= 310-340 grams/day), (9) low vegetables (TMREL= 280-320 grams/day), (10) low nuts and seeds (TMREL= 10-19 grams/day) (11) low legumes TMREL= 90-100 g/day, (12) low whole grains (TMREL= 140-160 g/day), (13) low calcium (TMREL=1.06 – 1.11 g/day), (14) low fibre (21-22 g/day), and low (15) polyunsaturated fatty acids (TMREL= 7-9% total energy intake).

GBD researchers ascribe another 27 million noncommunicable disease deaths to high levels of metabolic risk factors high BMI, high fasting plasma glucose (FPG), high low density lipoprotein cholesterol (LDL-C), and high systolic blood pressure (SBP)). Low levels of these metabolic risk factors are not also included as risk factors for NCD. For example, while the TMREL for BMI is 20 - 25 kg/M^2^, no population attributable risk for BMI < 20 is reported.^1^

This paper will use population weighted, formatted worldwide GBD data to begin to assess the impact of dietary and metabolic risk factors on noncommunicable disease deaths/100k/year (NCD). Findings will be compared with those derived from systematic literature reviews.

## Methods

As volunteer collaborators with the IHME, we utilise raw GBD ecological data (≈1.4 Gigabytes) on NCD, metabolic risk factors, dietary risk factors and covariates, and other risk factors of male and female cohorts 15-49 years old and 50-69 years old from each year 1990-2017 from 195 countries and 365 subnational locations (n=1120 cohorts). We also use 2019 GBD data on the dietary risks composite metric correlated with NCD. To maximally utilise the available GBD data, we average the values for ages 15-49 years old together with 50-69 years old for NCD and each risk factor exposure for each male and female cohort for each year. Finally, for each male and female cohort, data from all 28 years (1990-2017) on mean NCD and on each of the risk factor exposures are averaged using the computer software program R.

Food risk factors come as g/day consumed percapita. GBD dietary covariate data originally come from Food and Agriculture Organization surveys of animal and plant food commodities available percapita in countries worldwide—as opposed to consumed per capita.^2^ Supplementary Table 1 lists the relevant GBD risk factors, covariates, and other available variables with definitions of those risk factor exposures.^3^

GBD worldwide citations of over 12,000 surveys constituting ecological data inputs for this analysis are available online from IHME.^2^ The main characteristics of IHME GBD data sources, the protocol for the GBD study, and all risk factor values have been published by IHME GBD data researchers and discussed elsewhere.^4-7^ These include detailed descriptions of categories of input data, potentially important biases, and methodologies of analysis. We do not clean or pre-process any of the GBD data. GBD cohort risk factor and NCD data from the IHME have no missing records. Because the GBD data for the study come from IHME, no ethics committee approval or institutional review board review is needed for this post hoc statistical analysis. The updated 2019 raw data with all the variables from the GBD 2017 data we use for this analysis may be obtained by volunteer researchers collaborating with IHME.^8^

To weigh the country and subnational data according to population, internet searches (mostly Wikipedia) yields the most recent population estimates for countries and subnational states, provinces, and regions. The 1120 GBD cohorts available are population weighted by a software program in R into an analysis dataset with 7886 population weighted cohorts representing about 7.8 billion people in 2020. Each male or female cohort in the population-weighted analysis dataset represents approximately 1 million people (range: < 100,000 to 1.5 million). For example, India with about 1.234 billion people has 617 rows of the same data for males and 617 rows for females. Maldives, with about 445,000 people, has a single row of data for males and another for females. Without population-weighting the data, cohorts in India and Maldives each would have had one row for males and one row for females in the analysis dataset, invalidating the analysis results. World population data from the World Bank or the Organisation for Economic Co-operation and Development could not be used because they do not include all 195 countries or any subnational data.

This report follows the STrengthening the Reporting of OBservational studies in Epidemiology (STROBE) guidelines for reporting global health estimates.^9^

Supplementary Table 2 details how omega-3 fatty acid g/day is converted to fish g/day using data on the omega-3 fatty acid content of frequently eaten fish from the National Institutes of Health Office of Dietary Supplements (USA).^10^ As shown in Supplementary Table 3, we convert all of the animal and plant food data, including alcohol and sugary beverage consumption, from g/day to kcal/day. For the g to kcal conversions, we use the Nutritionix track app,^11^ which tracks types and quantities of foods consumed. Saturated fatty acids risk factor (0-1 portion of the entire diet) is not available with GBD data from 2017, so GBD saturated fatty acids risk factor data from GBD 2016 is used. Polyunsaturated fatty acid and trans fatty acid GBD risk factor data from 2017 (0-1 portion of the entire diet) are also utilised. These fatty acid data expressed as 0-1 portion of the entire diet are converted to kcal/day by multiplying by the kcal/day available for each cohort.

The principle outcome variable is noncommunicable disease deaths/100k/year (NCD). NCD is a combination variable consisting of the deaths/100k/year of over 100 diseases including cancers, diabetes, and diseases of the cardiovascular, respiratory, hepatic, renal, and central nervous system.

### Statistical methods

To determine the strengths of the risk factor correlations with mean NCD of population weighted worldwide cohorts (7886 cohorts) or subgroups of cohorts (e.g., highest NCD and lowest NCD cohorts, etc.), we utilise Pearson correlation coefficients: r, 95% confidence intervals (95% CIs), and p values. The subgroup of worldwide data for a sensitivity analysis is the subnational data from the UK, USA, Mexico, and Japan (n=730 cohorts). As with the worldwide data analysis, we report NCD correlating with risk factors and NCD trend correlating with risk factor trends for this subgroup.

We use SAS and SAS Studio statistical software 9.4 (SAS Institute, Cary, NC) for the data analysis.

## Results

Table 1 shows that the dietary risks composite risk factor correlated with NCD determined that 27.1 / 1426 (1.9%) of worldwide NCD are attributable to dietary risks. The dietary risks composite risk factor is moderately strongly correlated with NCD (r=0.390, 95% CI: 0.371 to 0.408, p<0.0001). In comparison with the worldwide findings, the USA dietary risks related to NCD is higher while the total NCD is lower, giving much higher percentages of NCD from dietary risks (3.62%: 42.90 dietary risks related NCD / 1184 total NCD=0.0362). The correlation of the dietary risks composite with NCD is very strong compared with worldwide and the other regions in this table (r=0.937, 95% CI: 0.924 to 0.948, p<0.0001).

**Table 1.** Dietary risks related NCD compared with total NCD worldwide (n=7886 cohorts, representing about 7.5 billion people in 2020)

| Regions or countries | NCD in region or country | NCD r | SD | Min | Max | Dietary risks versus total NCD r | 95% CI | p | (Dietary risks related NCD / Total NCD) * 100 = % |
| --- | --- | --- | --- | --- | --- | --- | --- | --- | --- |
| **World** | Dietary risks related NCD | 27.07 | 22.92 | 2.6 | 220 |  |  |  |  |
| **World** | Total NCD | 1426 | 463 | 424 | 4321 | 0.390 | 0.371 to 0.408 | <.0001 | 1.9 |
| **USA** | Dietary risks related NCD | 42.90 | 19.46 | 14.17 | 98.5 |  |  |  |  |
| **USA** | Total NCD | 1184 | 316 | 697 | 2312 | 0.937 | 0.924 to 0.948 | <.0001 | 3.62 |
| **UK** | Dietary risks related NCD | 13.06 | 14.67 | 2.71 | 61.1 |  |  |  |  |
| **UK** | Total NCD | 1195 | 330 | 777 | 1823 | 0.610 | 0.428 to 0.740 | <.0001 | 1.09 |
| **African Sahel** | Dietary risks related NCD | 25.03 | 11.28 | 11.7 | 47.1 |  |  |  |  |
| **African Sahel** | Total NCD | 1580 | 340 | 1115 | 3229 | 0.565 | 0.441 to 0.666 | <.0001 | 1.58 |
| **Central and Eastern Europe** | Dietary risks related NCD | 50.24 | 42.14 | 8.79 | 220 |  |  |  |  |
| **Central and Eastern Europe** | Total NCD | 2067 | 827 | 875 | 3193 | 0.355 | 0.253 to 0.447 | <.0001 | 2.43 |

While the USA and the UK have similar rates of NCD (NCD=1184 and 1195, respectively), the USA has more than threefold more the dietary risks composite risk factor related to NCD (42.90 /13.06=3.27).

As noted in the IHME GBD risk factors landmark article, the African Sahel region has the lowest level of the dietary risks composite related disability adjusted life years (DALYs) in the world. However, the African Sahel has above the worldwide average number of NCD (1580 versus 1426 worldwide). The GBD risk factors landmark article noted Central and Eastern Europe as having among the highest DALYs in the world. However, the range of total NCD in the African Sahel (1115-3229) largely overlaps the range of total NCD in Central and Eastern Europe (875-3193).

Table 2a shows worldwide NCD correlated with metabolic risk factors. Mean BMI, mean FPG, and mean LDL-C all negatively correlate with NCD (r=-0.308, 95% CI: −0.328 to −0.288, p<0.0001, r=-0.141, 95% CI: −0.163 to −0.119, p<0.0001, r=-0.341, 95% CI: −0.360 to −0.321, p<0.0001, respectively). Only mean SBP worldwide positively correlates with NCD (r=-0.068, 95% CI: 0.046 to 0.090, p<0.0001). On the other hand, Table 2b shows that the literature review derived composite dietary risks related to NCD are positively correlated with all four metabolic risk factors.

**Table 2.**
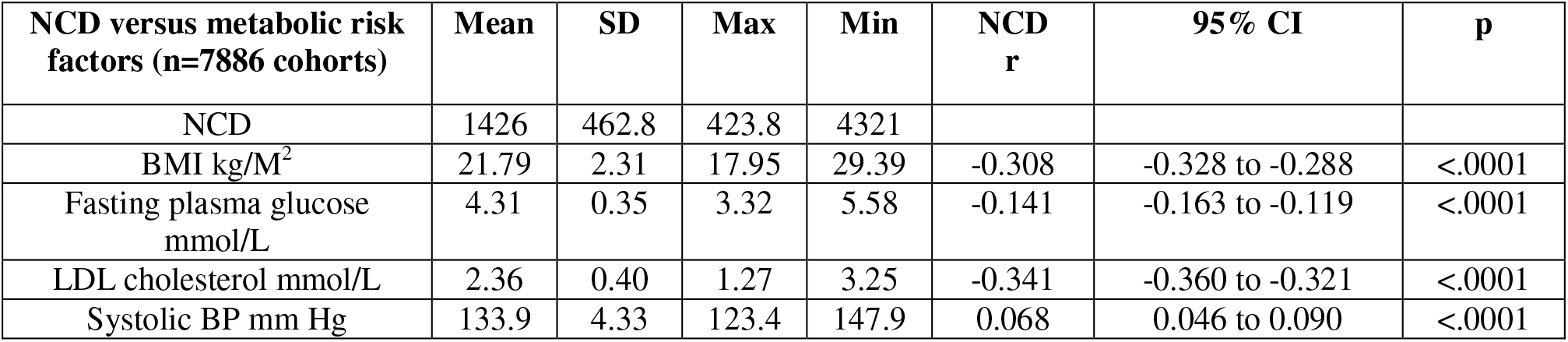
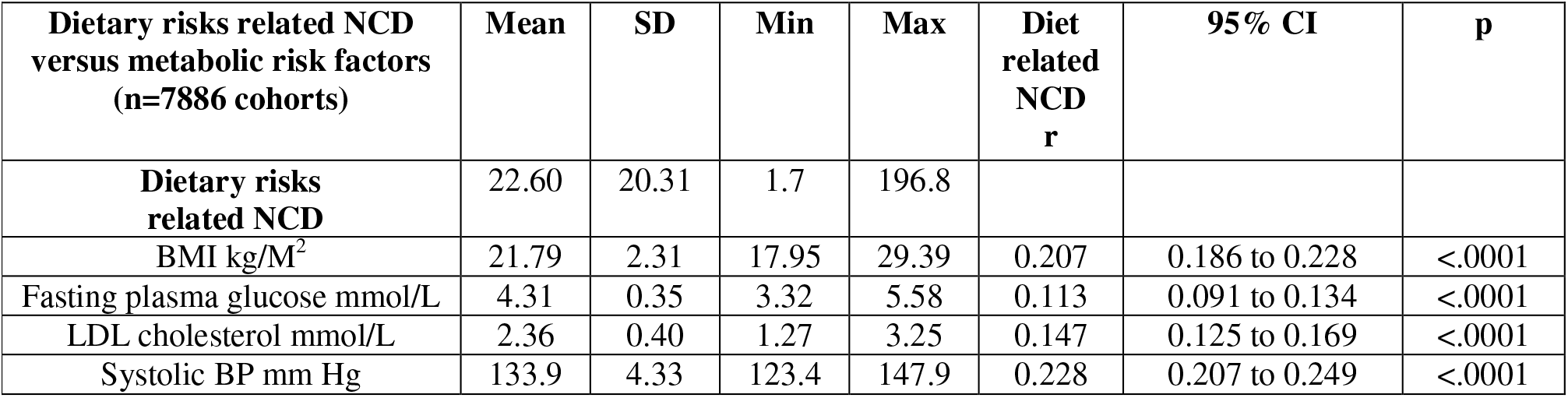
Worldwide NCD correlated with metabolic risk factors and dietary risks related NCD also correlated with metabolic risk factors. **Table 2a. Worldwide NCD correlated with metabolic risk factors** **Table 2b. Dietary risks related NCD correlated with metabolic risk factors**

Table 3 compares the NCD in cohorts with the lowest mean BMI (n=952 cohorts) with the NCD in the cohorts with the highest mean BMI (n=987 cohorts). The NCD is much higher in the low BMI cohorts (mean NCD in low BMI cohorts=1821 versus mean NCD in high BMI cohorts 1207). The low BMI group with high NCD had a small fraction of the animal food kcal/day of the high BMI group (low BMI six animal foods: 39.32 kcal/day versus high BMI six animal foods: 367.2) and much less intake of fatty acids (169.3 versus 492.9 kcal/day).

**Table 3.**
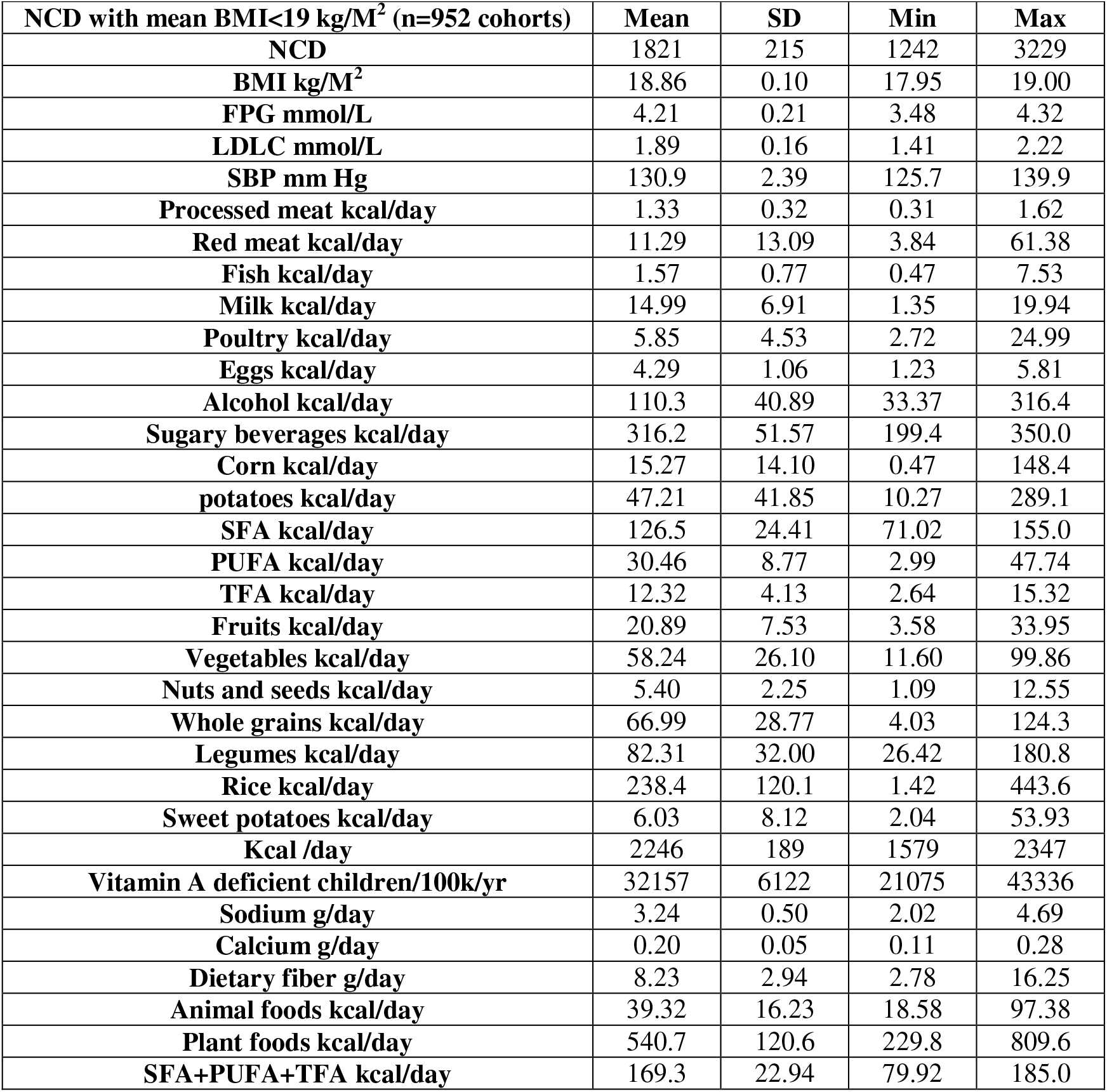
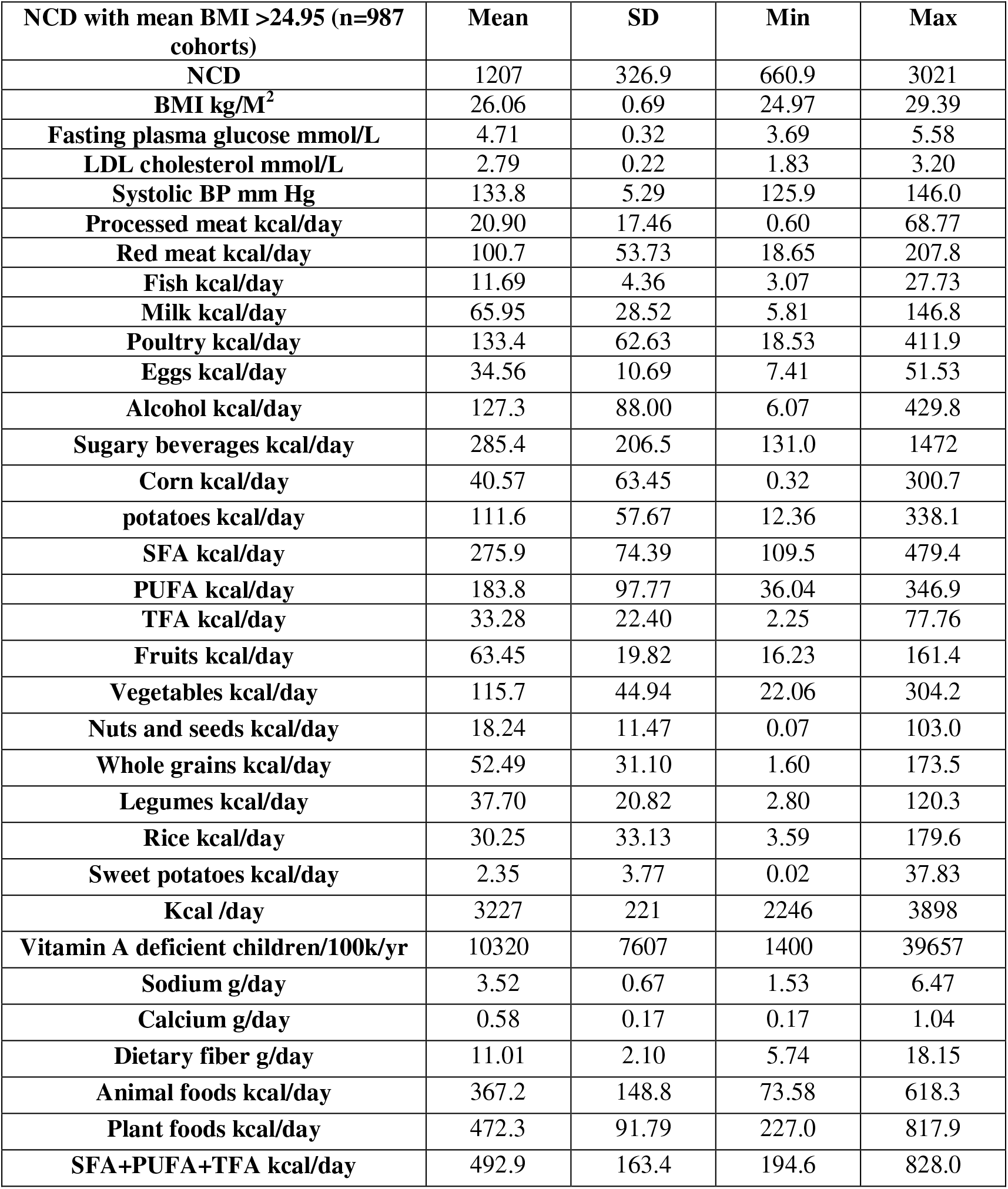
BMI<19.00 versus BMI >24.95 associated with NCD and NCD risk factors. **Table 3a. BMI<19 associated with NCD and NCD risk factors** **Table 3b. BMI>24.95 associated with NCD and NCD risk factors**

Table 4 compares cohorts with the lowest and highest rates of NCD—each group with about 1000 cohorts. The male and female NCD are averaged for each location because of the substantial differences in NCD between male and female cohorts. The mean animal food intake in the low NCD cohorts (animal foods=320.4 kcal/day) is over twice the worldwide average (animal foods=156.1 kcal/day) and nearly three times the mean animal food intake of the highest NCD cohorts (animal foods=117.3 kcal/day). SFA+PUFA+TFA is 66% higher in the lowest NCD cohorts compared with the highest NCD cohorts (406.3 kcal/day versus 244.5 kcal/day: 406.3/244.5=1.662). The rate of vitamin A deficiency in children is 75% higher in the highest NCD cohorts than the lowest NCD cohorts (24678 / 14082=1.752). Vitamin A is a fat soluble vitamin coming largely from animal foods.

**Table 4.**
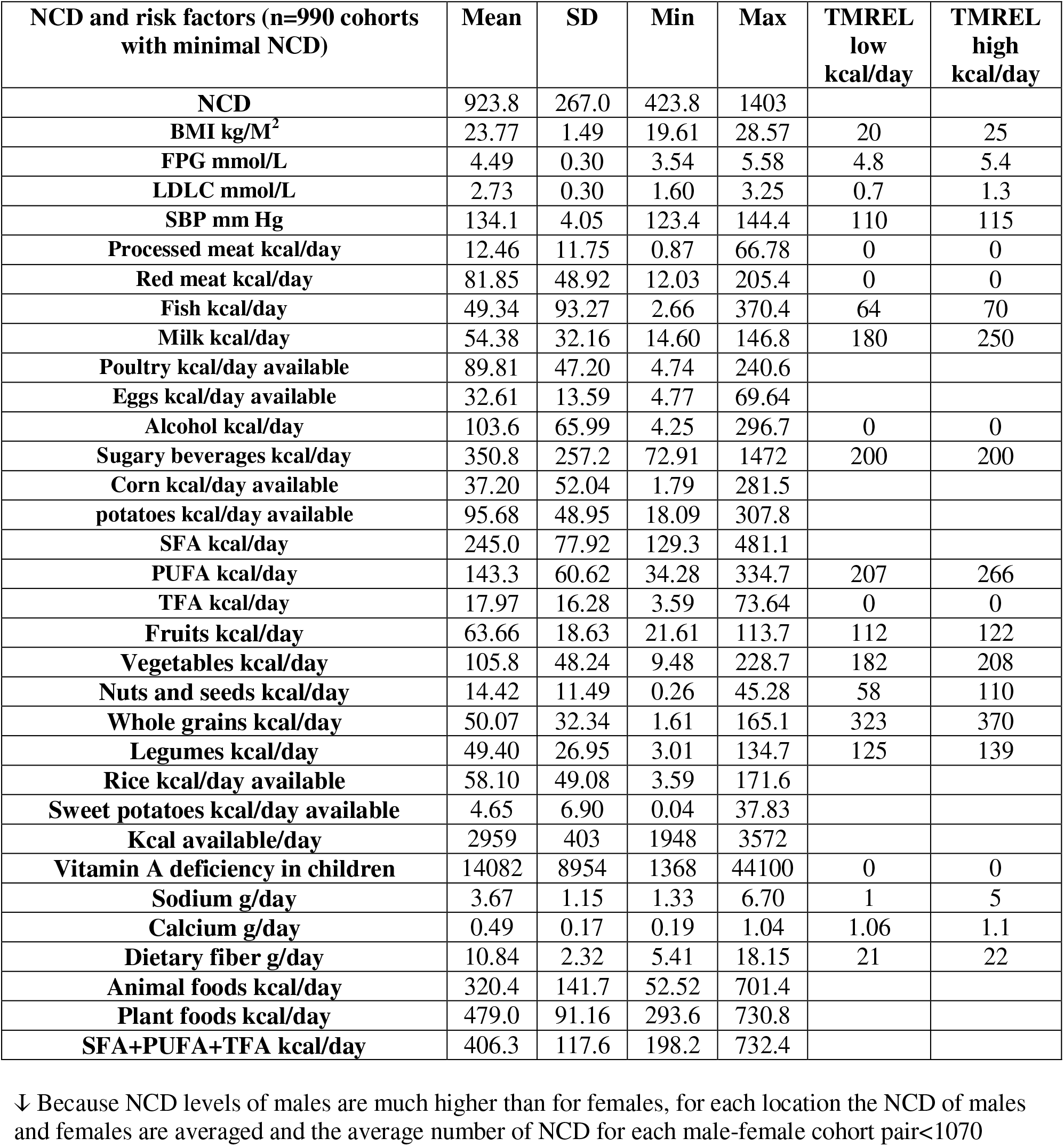
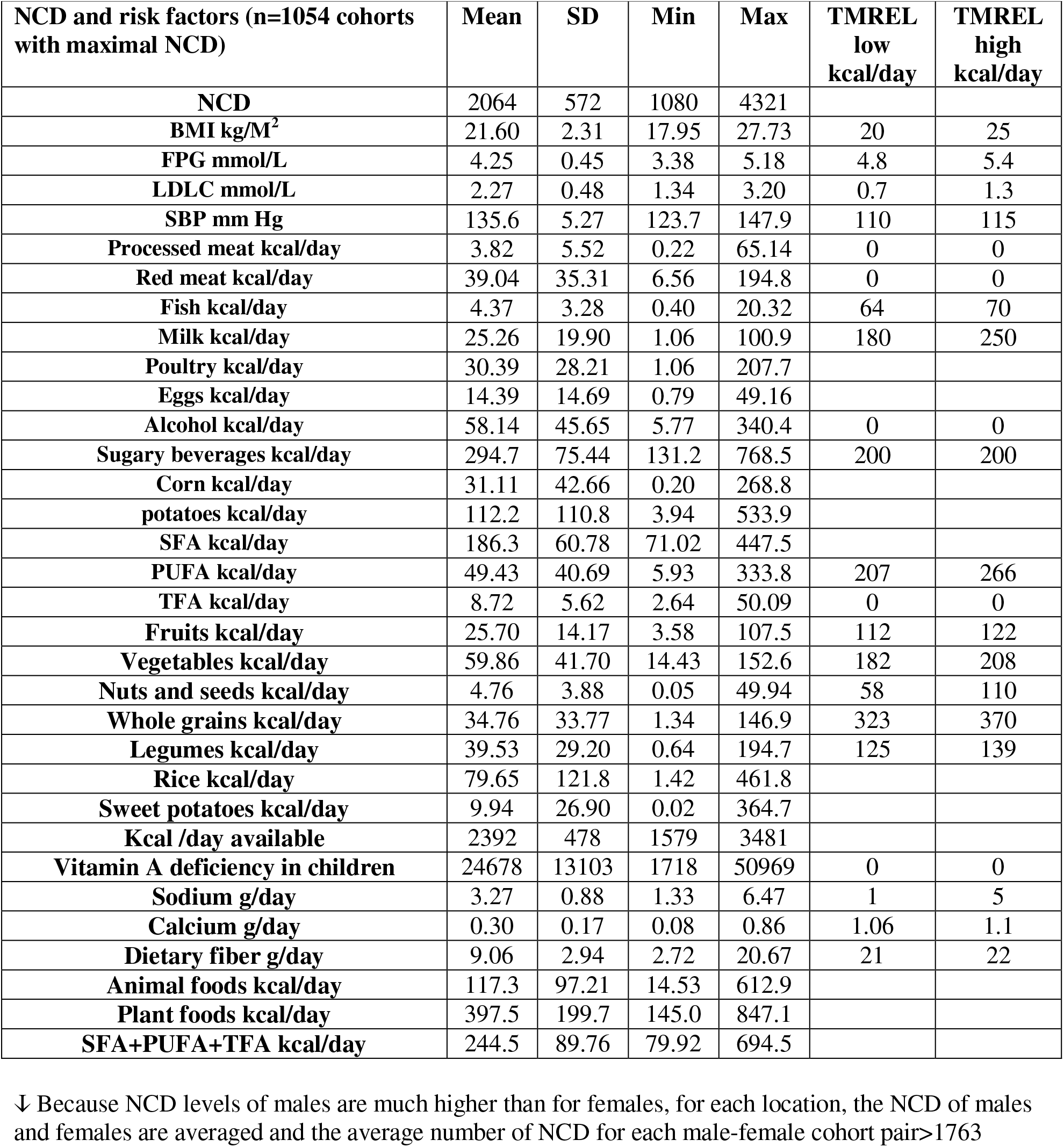
Comparing metabolic and dietary risk factor values in cohorts with the lowest and highest NCD. **Table 4a. Lowest NCD cohorts↓** **Table 4b. Highest NCD cohorts↓**

Table 4 also reports the TMREL levels of the metabolic risk factors, 15 dietary risk factors, alcohol, and vitamin A deficiency. The 990 cohorts with the lowest mean NCD of male-female pairs of cohorts have mean BMI=23.77, but the range goes from below to above the TMREL range (BMI range: 19.61 to 28.57). The 1054 cohorts with the highest NCD (mean NCD=2064) also have wide ranging BMI levels (mean BMI=21.60, range: 17.95 to 27.73). The 576 cohorts within the FPG TMREL range (FPG: 4.8 to 5.4 mmol/L) average 1409 NCD with a very wide range (NCD range: 426 to 3349). The 7307 cohorts with mean FPG < 4.8 mmol/L have mean NCD=1428, again with a very wide range (NCD range: 424 to 4321). Not a single cohort has a mean LDLC or mean SBP within the TMREL.

Table 5 shows the NCD trend (by least squared linear regression) from 1990 to 2017 correlated with the trends of metabolic and dietary risk factors. Notably, NCD trends downwards (−2.11 NCD) while all the metabolic risk factors and macronutrient risk factors trend up except for LDLC, corn availability, rice availability, sweet potato availability, and trans fatty acids. The NCD trend correlates negatively with BMI, FPG, SBP, and negatively with all macronutrient risk factors except for fish (p=0.86), sugary beverages, rice, sweet potatoes, and trans fatty acids.

**Table 5.** NCD trend versus risk factor trends.

| **NCD trend and risk factor trends worldwide (n=7886 cohorts)** | **Mean** | **SD** | **Min** | **Max** | **NCD trend v risk factor trend r** | **95% CI** | **p** |
| --- | --- | --- | --- | --- | --- | --- | --- |
| **NCD trend** | -2.11 | 46.54 | -119 | 355 |  |  |  |
| **BMI trend** | 0.08 | 0.03 | -0.02 | 0.22 | -0.055 | -0.077 to -0.033 | <.0001 |
| **FPG trend** | 0.02 | 0.01 | -0.01 | 0.04 | -0.158 | -0.179 to -0.136 | <.0001 |
| **LDLC trend** | 0.00 | 0.01 | -0.02 | 0.02 | 0.043 | 0.021 to 0.065 | 0.0001 |
| **SBP trend** | 0.02 | 0.11 | -0.48 | 0.31 | -0.055 | -0.077 to -0.033 | <.0001 |
| **Processed meat kcal/day trend** | 0.05 | 0.16 | -0.81 | 1.68 | -0.143 | -0.165 to -0.121 | <.0001 |
| **Red meat kcal/day trend** | 0.55 | 1.09 | -3.29 | 4.73 | -0.114 | -0.136 to -0.092 | <.0001 |
| **Fish kcal/day trend** | 0.22 | 0.95 | -0.13 | 9.79 | 0.002 | 0.020 to 0.024 | 0.8647 |
| **Milk kcal/day trend** | 0.19 | 0.30 | -2.02 | 2.50 | -0.072 | -0.094 to -0.050 | <.0001 |
| **Poultry kcal/day trend** | 1.15 | 1.04 | -0.87 | 7.66 | -0.020 | -0.042 to -0.002 | 0.0701 |
| **Eggs kcal/day trend** | 0.30 | 0.49 | -0.88 | 1.27 | -0.147 | -0.169 to -0.126 | <.0001 |
| **Alcohol kcal/day trend** | 0.03 | 0.16 | -0.91 | 1.99 | -0.270 | -0.290 to -0.249 | <.0001 |
| **Sugary beverages kcal/day trend** | 0.29 | 1.14 | -5.40 | 14.00 | 0.081 | 0.059 to 0.103 | <.0001 |
| **Corn kcal/day trend** | -0.03 | 0.60 | -4.36 | 2.81 | -0.056 | -0.078 to -0.034 | <.0001 |
| **Potatoes kcal/day trend** | 0.24 | 1.83 | -12.5 | 11.03 | -0.110 | -0.131 to -0.088 | <.0001 |
| **SFA trend** | 0.21 | 0.92 | -3.66 | 2.09 | -0.253 | -0.274 to -0.232 | <.0001 |
| **PUFA trend** | 1.00 | 0.96 | -3.03 | 5.47 | -0.026 | -0.048 to -0.004 | 0.0195 |
| **TFA trend** | -0.21 | 0.45 | -2.68 | 0.01 | 0.123 | 0.102 to 0.145 | <.0001 |
| **Fruits kcal/day trend** | 0.62 | 0.60 | -2.08 | 4.10 | -0.095 | -0.117 to -0.073 | <.0001 |
| **Vegetables kcal/day trend** | 1.39 | 1.61 | -7.96 | 8.47 | -0.027 | -0.049 to -0.005 | 0.0151 |
| **Nuts and seeds kcal/day trend** | 0.28 | 0.27 | -0.34 | 3.12 | -0.136 | -0.158 to -0.115 | <.0001 |
| **Whole grains kcal/day trend** | 0.17 | 0.58 | -2.60 | 8.46 | 0.162 | 0.141 to 0.183 | <.0001 |
| **Legumes kcal/day trend** | 0.37 | 0.98 | -3.24 | 5.40 | -0.087 | -0.109 to -0.065 | <.0001 |
| **Rice kcal/day trend** | -0.78 | 1.29 | -5.05 | 3.50 | 0.191 | 0.170 to 0.212 | <.0001 |
| **Sweet potatoes kcal/day trend** | -0.46 | 1.34 | -4.82 | 3.91 | 0.124 | 0.102 to 0.146 | <.0001 |
| **Kcal /day available trend** | 12.12 | 10.10 | -28.1 | 47.89 | -0.180 | -0.201 to -0.158 | <.0001 |
| **Vitamin A deficient children/100k/yr trend** | -269 | 253 | -1425 | 169.4 | -0.057 | -0.078 to -0.034 | <.0001 |
| **Sodium g/day trend** | -0.001 | 0.044 | -0.219 | 0.085 | -0.107 | -0.128 to -0.085 | <.0001 |
| **Calcium g/day trend** | 0.003 | 0.003 | -0.007 | 0.018 | 0.155 | 0.133 to 0.176 | <.0001 |
| **Dietary fiber g/day trend** | 0.06 | 0.07 | -0.23 | 0.54 | -0.073 | -0.095 to -0.051 | <.0001 |

As a contrast to the worldwide GBD data analysis, Table 6a shows NCD correlated with risk factors from subnational data of the USA, UK, Mexico, and Japan (n combined=730 cohorts). In these mostly developed countries (Mexico is classified advanced developing^12^) with NCD much lower than worldwide (NCD four countries=1071 versus worldwide=1426), mean BMI is strongly positively correlated with NCD (r=0.504, (95% CI 0.447 to 0.556, p<0.0001). As opposed to the worldwide NCD analysis in Table 4, processed meat, red meat, milk, poultry, SFA, PUFA, and kcal available are all at least moderately strongly correlated with NCD. As opposed to the worldwide NCD trend’s correlations in Table 5; the four country NCD trend is positively correlated with the trends of processed meat, red meat, fish, poultry, eggs, and SFA.

**Table 6.**
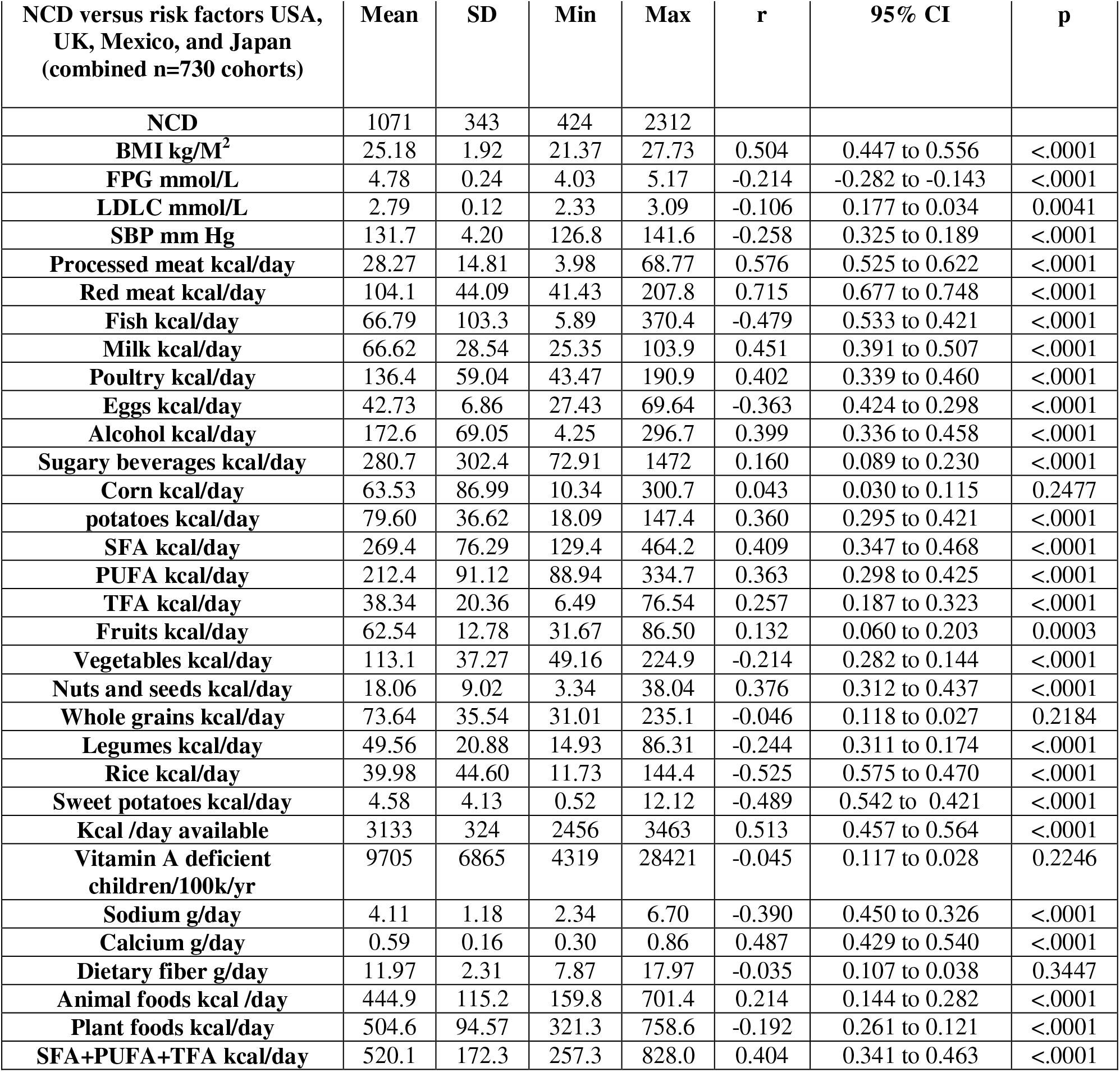
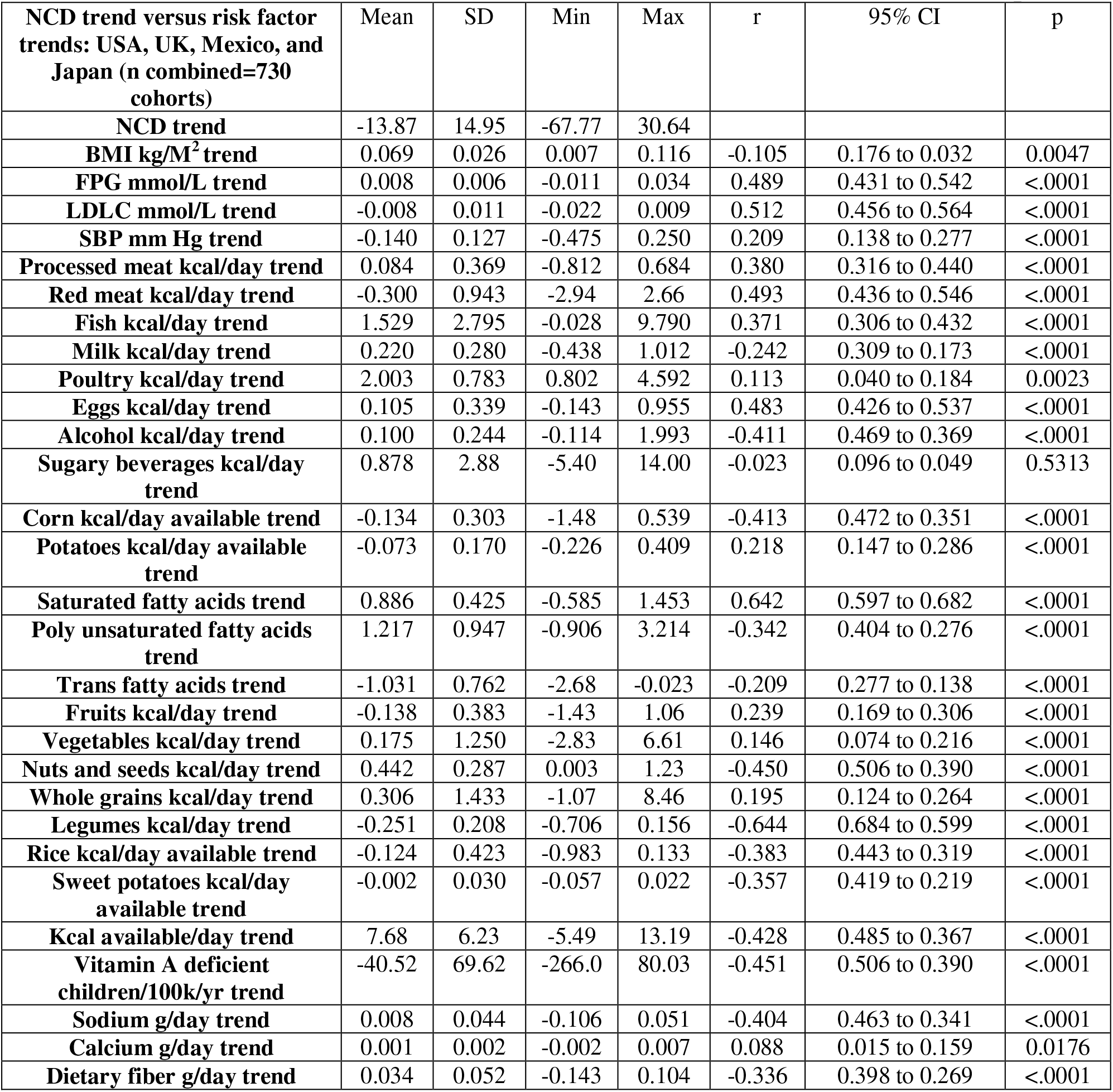
NCD and the NCD trend versus risk factors and risk factor trends in USA, UK, Mexico, and Japan combined. **Table 6a. NCD correlated with risk factors in USA, UK, Mexico, and Japan combined** **Table 6b. NCD trend correlated with risk factor trends in USA, UK, Mexico, and Japan**

## Discussion

Table 1 shows that systematic literature review derived dietary risks account for only 1.9%: (27.07 dietary risks related NCD / 1426 total NCD), bringing the dietary risks metric into question. Regarding the USA, Table 1 shows the relatively high percent of NCD deaths accounted for by dietary risks (% NCD due to dietary risks: 3.62%). Additionally, the USA has a relatively strong correlation of dietary risks correlated with NCD with total NCD (r=0.937, 95% CI: 0.924 to 0.948, p<0.0001). This suggests that the dietary risks metric, based on systematic literature reviews, may be biased toward the NCD data from the USA and other developed countries over developing countries.

The Sahel region of Africa, which has the world’s lowest shares of DALYs attributed to dietary risks of any region,^1^ has higher than the worldwide average of NCD (Sahel region NCD: 1580 versus worldwide NCD: 1426. This also implies that the dietary risks metric may be biased toward dietary risk factors as reported in medical literature from developed countries. In Central and Eastern Europe, which literature reviews show have among the world’s highest shares of DALYs attributed to dietary risks of any region,^1^ the much higher than average NCD has a range that greatly overlaps the range of the Sahel region of Africa (Central and Eastern Europe: mean NCD=2067, range: 875-3193 versus Sahel region: mean NCD=1580, range: 1115-3229). This comparison further calls into question the evidence basis of the composite dietary risks metric.

In Tables 2a and 2b, NCD negatively correlate with BMI, FPG, and LDLC. However, literature review derived dietary risks related to NCD positively correlate with BMI, FPG, and LDLC. This also suggests that the dietary risks metric is biased toward the survey findings of the USA and other developed countries in which high levels of BMI, FPG, and LDLC are more prevalent than in developing countries.

Tables 3a and 3b detail how NCD in the cohorts lowest in BMI are much higher than in cohorts highest in BMI (BMI<19 kg/M^2^: mean NCD=1821 versus BMI >24.95 kg/M^2^: mean NCD=1207). This is further suggestive evidence that literature derived high BMI, high FPG, and high LDLC attributions of NCD and noncommunicable disease DALYs are biased toward developed countries over developing countries.

Table 3 also shows that the high NCD of cohorts with large numbers of underweight cohorts (BMI< 18.5) are associated with very low mean animal food kcal/day intake that is about one-ninth the animal food kcal/day intake of the highest BMI cohorts (mean animal food intake lowest mean BMI cohorts=39.32 kcal/day versus mean animal food intake of the cohorts with the highest mean BMI=367.2 kcal/day: 367.2/39.32=9.33). These worldwide data are at variance with the lipid hypothesis (saying that dietary saturated fat and cholesterol, largely from animal foods, cause high LDLC and atherosclerosis) as the dominant dietary risk factor for cardiovascular disease (CVD). However, as is shown in Table 6, dietary SFA and cholesterol may still strongly contribute to CVD mostly in developed countries with high intakes of animal foods and the associated high rates of overweight and obesity.

Compared with analysing these worldwide GBD data, systematic literature reviews with individual participant analysis would likely be much better than using GBD mean cohort values of with about 1 million people per cohort in documenting CVD and NCD associated with obesity, overweight, normal weight and underweight. Flegal, et. al., with an individual participant level systematic review of BMI levels related to longevity, report that, compared to normal weight (18.5≤BMI<25), the summary HRs are the following: overweight (25≤BMI<30): (HR r=0.94 (95% CI, 0.91-0.96), stage 1 obesity (30≥BMI<35): HR r=0.95 (95% CI, 0.88-1.01), and above stage 1 obesity (BMI≥35): HR r=1.29 (95% CI, 1.18-1.41).^13^ A systematic review by Aune, et. al. including underweight people finds BMI=20 gives a 15% increase in all cause mortality (HR r=1.15 (95% CI, 1.13-1.17), p<0.0001) and BMI=17.5 gives a 47% increase in all cause mortality (HR r=1.47 (95% CI, 1.43-1.51), p<0.0001)^14^ These are consistent with the Table 3 findings for low BMI.

While GBD data on mean BMIs of cohorts cannot probe the impact of overweight and obesity on NCD as can literature reviews, GBD data should be very informative for estimating the NCD impact of cohorts with high BMI on average (i.e., mean BMI ≥25) in comparison with cohorts that are underweight and normal weight on average (i.e., mean BMI <25). The 973 cohorts with mean BMI ≥25 have mean NCD=1208, while the 6913 cohorts with mean BMI<25 have average NCD=1457. The BMI≥25 cohorts account for 10.5% of the total NCD and the BMI<25 account for the remaining 89.5% of the NCD (BMI≥25 cohorts: 973 cohorts * 1208 (mean NCD) = 1,175,384 total NCD versus BMI<25 cohorts: 6913 cohorts * 1457 (mean NCD) = 10,072,241 total NCD: 1,175,384 + 10,072,241 =11,247,625 (total NCD), mean BMI ≥25: 1,175,384 / 11,247,625 = 0.1045 and mean BMI <25: 10,072,241 / 11,247,625 = 0.8955).

BMI≥25 cohorts also have higher FPG and LDLC: FPG mean=4.71 and LDLC mean=2.79 versus the BMI<25 cohorts with: FPG mean=4.25 and LDLC mean=2.30. The mean levels of SBP of the BMI≥25 cohorts and the BMI<25 cohorts differ by only 0.2 mm Hg (BMI≥25 cohorts SBP mean=133.71 versus BMI<25 cohorts SBP mean=133.91). These comparisons strongly suggest that high levels of dietary and metabolic risk factors should account for about 10% of the worldwide NCD rather than about 62%, which can be derived from what is reported from the analyses based on systematic literature reviews ((35 million worldwide deaths/year attributable to dietary risks and metabolic risk factors / 56.5 million worldwide deaths/year^1^) =0.6195).

These GBD data showing that low consumption of animal foods and fatty acids correlate strongly with low BMI and high NCD suggests that deficiencies of the fat soluble vitamins A, D, E, and K might contribute to CVD and thereby NCD. Animal foods are the primary sources of each of these vitamins and their absorption in the gut depends on adequate fatty acid intake that also comes largely from animals. By different mechanisms, deficiencies of each of these fat soluble vitamins raises the risk of CVD (vitamins A,^15^ D,^16^ E,^15^ and K^17^).

Our “fat soluble vitamin hypothesis” is that the high rates of CVD and, consequently, NCD in countries with low animal food and low fatty acid intake stem to a significant degree from deficiencies of fat soluble vitamins. For instance, over three times the vitamin A deficiency prevalence in children occurs in the low BMI cohorts compared to high BMI cohorts (32.2% of children < 5 years old of low BMI cohorts have vitamin A deficiency versus 10. 3% of children from the high BMI cohorts, see Table 3).

Table 4, which compares 990 cohorts with the lowest NCD with 1054 cohorts with the highest NCD, gives similar findings as Table 3 with low and high BMI levels. The metabolic and dietary risk factors for the mean NCD of the lowest 1054 cohorts does not perceptively correspond better with TMREL ranges than do the mean NCD of the highest 990 cohorts.

Again the higher intake of animal foods is associated with lower levels of NCD in Table 4. This is all consistent with the hypothesis that fat soluble vitamin deficiencies significantly contribute to NCD. The 75% higher rate of vitamin A deficiencies of the highest cohorts in NCD (vitamin A deficiency rate =24,687/100k/year) versus the lowest NCD cohorts (14,082/100k/year) and is another example.

Table 5 shows how the worldwide downward NCD trend from 1990 to 2017 relates to the trends of the metabolic and dietary risk factors. While NCD trends downwards by −2.11 /year on average, BMI, FPG, and SBP are trending upwards and LDLC is neutral. The NCD trend negatively correlating with the BMI and FPG trends reinforces Table 2a data showing negative correlations of NCD with BMI and FPG.

These metabolic risk factor correlations with NCD and the metabolic risk factor trends with the NCD trend do not exclude that high levels of metabolic risk factors contribute significantly to NCD. We have a vast literature relating individuals with high metabolic risk factor levels with increased NCD. However, these GBD cohort data, now including the trend analysis, suggest that BMI, FPG and perhaps LDLC correlate negatively with NCD.

In Table 5, all of the 20 food commodities are trending upwards except for corn, trans fatty acids, rice and sweet potatoes. Kilocalories (kcal) /day available, a covariate, goes up by12 kcal/day/year. As might be expected, trends of sugary beverages and trans fatty acid (the only clearly always harmful food risk factors) positively correlate with the NCD trend. With worldwide development, vitamin A deficiency in children is on the decline (−269 cases/100k/year from 1990-2017), most likely related to increased animal foods, which contain most of the dietary vitamin A and fatty acids, which are required for gut absorption of vitamin A.

Despite the major increase in percapita food intake worldwide from 1990 to 2017, the NCD trend and the trends of most macronutrients are still negatively correlated, indicating that more food percapita is needed to drive down NCD, especially in developing countries. Dietary choices need to be improved in places, especially developed countries, where abundant food availability allows for and often promotes high levels of metabolic risk factors.

Table 4 shows that the cohorts with minimal NCD had higher mean sodium intake (3.67 g/day) than the cohorts with maximum NCD (3.27 g/day). Accordingly, sodium g/day trend correlates negatively with the NCD trend. This suggests that sodium lowering public health campaigns may not be effective to benefit population health. However, individuals with CVD, hypertension, or chronic diseases causing fluid overload still should reduce dietary sodium.

Table 6 shows how using data primarily from developed countries can easily support the lipid hypothesis that defines saturated fat and cholesterol as the major dietary drivers of CVD and NCD. However Table 6 findings are not generalisable to the world. In the four countries with subnational GBD data in Table 6, the high level animal food consumption (animal foods=445 kcal/day) reverses the worldwide negative correlation of animal food with NCD (Four countries cohorts: NCD versus animal foods, r=0.214, 95% CI (0.144 to 0.282), p<0.0001). Meat is especially strongly positively correlated with NCD (NCD versus processed meats r=0.576, 95% CI (0.525 to 0.622), p<0001 and NCD versus red meat r=0.715, 95% CI (0.677 to 0.748), p<0001).

The unexpected positive correlations of NCD with corn availability (covariate), potato availability (covariate) and fruit in Table 6 may be explained. Corn availability includes high fructose corn syrup used widely in developed countries. Corn availability and sugary beverages are highly positively correlated in these cohorts (corn availability versus sugary beverages, r=0.944, 95% CI 0.935 to 0.951, p<0.0001). Fruits include 100% fruit juice in the GBD database (see Supplementary Table 1). In the USA, about one-third of fruits consumed are in the form of 100% fruit juices.^18^ In these four countries combined, fruits are positively correlated with sugary beverages (fruits versus sugary beverages, r=0.186, 95% CI 0.115 to 0.255, p<0.0001). Potato availability includes highly processed potatoes like potato chips and French fries. According to the International Potato Center, at least 50% of potatoes consumed worldwide are highly processed.^19^ Processed potatoes may contain up to 6 g (or 54 kcal) per 100 kcal serving.^20^ While SFA worldwide averages only 191.3 kcal/day and is negatively correlated with NCD (SFA versus NCD worldwide (n=7886 cohorts), r=-0.253, 95% CI −0.274 to −0.232, p<0.0001), SFA averages 269.4 kcal/day and is positively correlated with NCD in these four mostly developed countries combined (SFA versus NCD four countries with subnational data (n=730 cohorts), r=0.642, 95% CI 0.597 to 0.682, p<0.0001). The lipid hypothesis is strongly supported by data on people from these four countries and other developed countries.

As mentioned in the IHME GBD 2019 risk factor paper, the assessment of the joint effects of dietary and other risk factors depends on two critical factors: the correlation of risk exposure and the estimation of the joint effects of groups of risks together.^1^ While the estimates of exposure to the individual dietary risk factors appears excellent and improving with each iteration of the GBD project, the estimation of the joint effects of the 15 food variables composite dietary risks and the metabolic risk factors are problematic, as discussed regarding Tables 1 and 2 above.

The IHME GBD methodology assumption that, for each age-sex-location-year, the estimates of the prevalence of exposure of each of the dietary variables are independent from each other is said to be supported by findings from USA data from the National Health and Nutrition Examination Survey. While this may have been the case for the USA where intakes of dietary commodities vary much less than they do worldwide, the assumption of independence of exposures of different dietary variables worldwide is highly dubious. As shown in Tables 3 and 4, there are marked differences between animal foods intake (four dietary risk factors and two dietary covariates) between cohorts with the lowest and highest mean BMI and the lowest and highest mean NCD. The GBD 2019 risk factor paper states in the limitations,

> “To avoid over-estimation of the joint effects, we computed the non-mediated relative risks and then assumed that non-mediated relative risks are multiplicative. This approach does not capture potential synergy between relative risks in which some combinations might be super-multiplicative. For some areas such as diet, the joint estimation is very important for public policy. Further, more detailed work is needed to strengthen the evidence base for understanding mediation.”^1^

This GBD formatted data analysis is perhaps the detailed work that is needed to strengthen the evidence base for understanding mediation and for more accurately determining the deaths and DALYs attributable to dietary and other risk factors.

Also mentioned under limitations in the GBD risk factor paper, “…we assume that relative risks as a function of exposure are universal and apply in all locations and time periods.”^1^ Tables 4-6 of this GBD data analysis gives a clear example of how the level of exposure of animal foods in cohorts worldwide versus in the four countries with subnational data lead to diametrically opposing correlations between NCD and the six animal foods kcal/day. Likewise, Table 6 shows the worldwide NCD trend negative correlations with trends of animal foods and SFA are reversed in the subnational data analysis (n=730 cohorts) for processed meat, red meat, milk, poultry and SFA.

## Limitations

The GBD data on animal foods, plant foods, alcohol, sugary beverages, and fatty acids are not comprehensive and comprised only 1191.4 kcal/day on average worldwide. Subnational data are available on only four countries. Because the data formatting and statistical methodology are new, this is necessarily a post hoc analysis and no pre-analysis protocol is possible. This GBD data analysis should be repeated with the most recently released GBD 2019 data when it becomes available to volunteer collaborators.

### Generalisability

The new methodology presented for probing the GBD raw data to find correlations between risk factors (e.g., dietary and metabolic risk factors) and health outcomes (e.g., NCD) can be applied to any of the risk factors and health outcomes available with the new GBD 2019. The intake of each of the 20 dietary variables in this analysis that corresponds with the 990 cohorts with the lowest NCD worldwide will not be the optimal healthy diet for everyone. The total kcal/day of animal foods, plant foods and fatty acids are all important. Having comprehensive data on dietary inputs is key to more accurate and reliable analyses. Baseline metabolic risk factor levels are important. The optimal diet to lower NCD risk for someone with BMI=35 will be different than in someone with BMI=20. This analysis is merely a proof of concept the the GBD data analysis methodology can derive plausible results and that the GBD data analysis methodology can be complimentary to the systematic literature review methodology.

## Conclusion

Statistically analysing GBD data provides a tool to compliment the systematic literature review methodology in forming policy recommendations, clinical practice guidelines, and public health recommendations. Eventually, GBD data analysis can bring more rigor, precision, and consensus to the field of population health, especially in the areas of dietary and metabolic risk factors.

## Supporting information

Strobe checklist

## Data Availability

The raw, unformatted data used in this analysis is now out of date. The 2019 GBD data on all the variables in this analysis may be obtained from the IHME by volunteer collaborating researchers. Our data formatting software code in R and SAS and our formatted database are available on request to researchers.

---

**Supplementary Table 1.** Definitions of IHME GBD risk factors and covariates related to BMI.

| Variables | Definition |
| --- | --- |
| **Alcohol** | Any alcohol consumption (g/day) |
| **Body-mass index** | Body mass index (BMI) ($\text{kg}/\text{m}^2$)—the dependent variable of interest |
| **Corn** | Corn availability percapita (g/day), a covariate |
| **Eggs** | Eggs availability percapita (g/day) a covariate |
| **Fasting plasma glucose** | Fasting plasma glucose (mmol/L) |
| **Fish** | This variable expressed in g/day is derived by determining the weight of fish in g corresponding to 1 g of omega-3 fatty acids (eicosapentaenoic acid and docosahexaenoic acid) by averaging the fish g per 1 g of omega-3 fatty acids 20 species of fish= 117.04 g/day fish/1 g/day omega-3 fatty acids (Supplementary Table 2) |
| **Fruits** | Consumption of fruits (includes fresh, frozen, cooked, canned, or dried fruit but excludes fruit juices and salted or pickled fruits) (g/day) |
| **Kilocalories available /day** | The mean number of kilocalories percapita available per day to people in each location (kcal/day available), a covariate |
| **LDL cholesterol** | Serum low-density lipoprotein cholesterol (mmol/L) |
| **Legumes** | Consumption of beans, lentils, pulses (g/day) |
| **Milk** | Consumption of milk including non-fat, low-fat, and full-fat milk but excluding soy milk and other plant derivatives (g/day) |
| **Nuts and seeds** | Consumption of nuts and seeds (g/day) |
| **Poultry** | Poultry availability percapita (g/day), a covariate |
| **Potatoes** | Potatoes availability percapita (g/day), a covariate |
| **Processed meat** | Consumption of any processed meat (includes meat preserved by smoking, curing, salting, or addition of chemical preservatives, including bacon, salami, sausages, or deli or luncheon meats like ham, turkey, and pastrami) (g/day) |
| **Red meat** | Consumption of red meat (includes beef, pork, lamb, and goat but excludes poultry, fish, eggs, and all processed meats) (g/day) |
| **Rice** | Rice availability percapita (g/day), a covariate |
| **Seafood omega-3 fatty acids** | Seafood omega-3 fatty acids (eicosapentaenoic acid and docosahexaenoic acid) in tablet or fish form (g/day) |
| **Sugar-sweetened beverages** | Consumption of any beverage with $\geq 50$ calories of sugar per one-cup serving, including carbonated beverages, sodas, energy drinks, fruit drinks but excluding 100% fruit and vegetable juices (g/day) |
| **Sweet potatoes** | Sweet potato availability percapita (g/day), a covariate |
| **Systolic blood pressure** | Systolic blood pressure (mm Hg) |
| **Vegetables** | Consumption of frozen, cooked, canned, or dried vegetables (excluding legumes salted or pickled, juices, nuts and seeds, and starchy vegetables such as potatoes or corn) (g/day) |
| **Whole grains** | Consumption of whole grains (bran, germ, and endosperm in their natural proportions) from breakfast cereals, bread, rice, pasta, biscuits, muffins, tortillas, pancakes, and others (g/day) |

**Supplementary Table 2.** Omega-3 Fatty Acid g to fish g calculation¶.

| **Fish** | **DHA**<br>**g/3**<br>**ounce**<br>**fish** | **EPA**<br>**g/3**<br>**ounce**<br>**fish** | **Omega-3 Fatty**<br>**Acids (DHA +**<br>**EPA) g/3 ounce**<br>**fish mean** | **Fish 3**<br>**ounces**<br>**= 85.02**<br>**g** | **Fish (g) per**<br>**omega-3**<br>**Fatty Acids**<br>**(g)=columns**<br>**E/F** |
| --- | --- | --- | --- | --- | --- |
| **Salmon Atlantic farmed** | 1.24 | 0.59 |  |  |  |
| **Salmon Atlantic wild** | 1.22 | 0.35 |  |  |  |
| **Herring Atlantic** | 0.94 | 0.77 |  |  |  |
| **Sardines canned in tomato**<br>**sauce drained** | 0.74 | 0.45 |  |  |  |
| **Mackerel Atlantic** | 0.59 | 0.43 |  |  |  |
| **Salmon pink canned**<br>**drained** | 0.63 | 0.28 |  |  |  |
| **Trout rainbow wild** | 0.44 | 0.40 |  |  |  |
| **Oysters eastern wild** | 0.23 | 0.30 |  |  |  |
| **Sea bass** | 0.47 | 0.18 |  |  |  |
| **Shrimp** | 0.12 | 0.12 |  |  |  |
| **Lobster** | 0.07 | 0.10 |  |  |  |
| **Tuna light canned in water**<br>**drained** | 0.17 | 0.02 |  |  |  |
| **Tilapia** | 0.11 |  |  |  |  |
| **Scallops** | 0.09 | 0.06 |  |  |  |
| **Cod Pacific** | 0.1 | 0.04 |  |  |  |
| **Tuna yellowfin** | 0.09 | 0.01 |  |  |  |
| **Mean DHA and EPA**<br>**Omega-3 Fatty Acids g/3**<br>**ounce fish** | 0.4531 | 0.2733 |  |  |  |
| **Calculations total Omega-3**<br>**FA g to fish g** |  |  | 0.7264 | 85.02 | 117.043 |

¶¶ Data on omega-3 fatty acid content of varieties of fish came from the National Institutes of Health Office of Dietary Supplements (USA)

**Supplementary Table 3.**
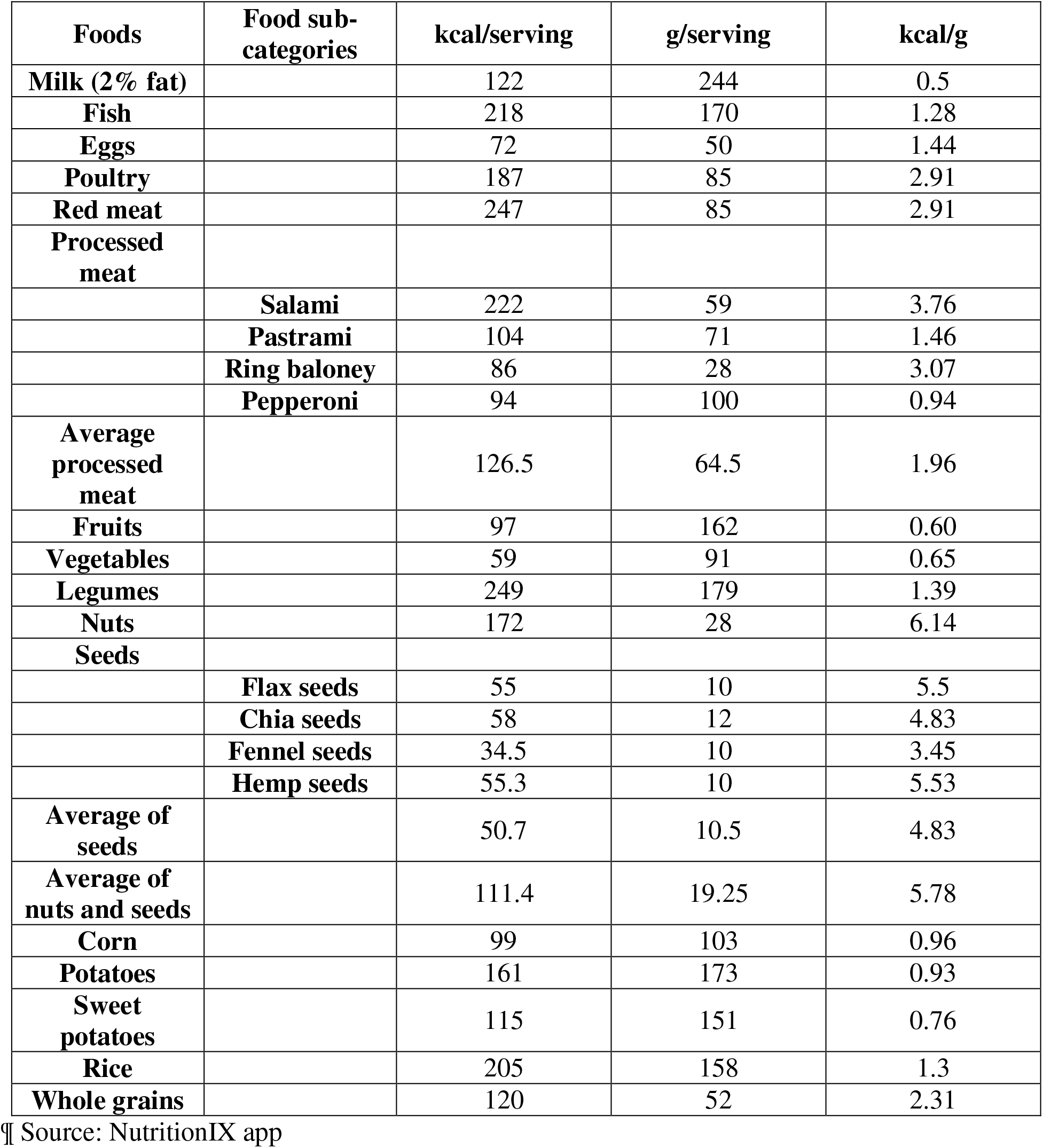
Calculations of kcal/day from g/day of animal and plant foods¶.

## Authors’ contributions

DKC acts as guarantor; conceived and designed the study, acquired and analysed the data, interpreted the study findings, drafted the manuscript, critically reviewed and edited the manuscript and tables, and approved the final version for publication.

CW designed software programs in R to format and population weight the data, aided with the SAS statistical analysis, critically reviewed the manuscript, and approved the final version for publication.

The corresponding author attests that all listed authors meet authorship criteria and that no others meeting the criteria have been omitted.

## Copyright

The authors retain the copyright to the paper

The Patient and Public Involvement

- When and how were patients/public first involved in the research? IHME acquired, catalogued and extracted information from over 12,000 surveys from government and non government agencies in order to compile the GBD database.
- How were the research question(s) developed and informed by their priorities, experience, and preferences? The diverse surveys contributing to the GBD database had many different priorities and intentions unrelated to this post hoc analysis. The current analysis research questions have been developed to give researchers, policymakers, and the public the methodological tool to quantify the impacts of dietary and other risk factor patterns.
- How were patients/public involved in
  a. the design and conduct of the study? The way patients/public were involved in the collection of the surveys has not been systematically studied and reported on. Patients/public are the intended beneficiaries of this analysis of the GBD data.
  b. choice of outcome measures? The outcome measures chosen all relate to human health.
  c. recruitment to the study? The recruitment methods vary by the survey.
- How were (or will) patients/ public be involved in choosing the methods and agreeing plans for dissemination of the study results to participants and linked communities? IHME and all the health surveyors that contributed will make these decisions. This analysis of the data will be available to all by open access.

## Competing interests statement

Both authors have completed the ICMJE uniform disclosure form at www.icmje.org/coi_disclosure.pdf and declare: no support from any organisation for the submitted work; no financial relationships with any organisations that might have an interest in the submitted work in the previous three years; no other relationships or activities that could appear to have influenced the submitted work.

## Contributors

Martin Sebera, from the Department of Kinesiology, Faculty of Sports Studies Masaryk University, Czech Republic, critiqued statistical aspects of the manuscript and provided useful input. Pavel Grasgruber, from Masaryk University, Czech Republic, provided suggestions after reviewing the manuscript.

## Transparency declaration

The manuscript is an honest, accurate, and transparent account of the study being reported; that no important aspects of the study have been omitted; and that any discrepancies from the study as planned have been explained.

## Ethics

Studies based solely on data from IHME GBD database do not need approval from any bioethics committee.

## Funding

This research received no specific grant from any funding agency in the public, commercial or not-for-profit sectors. The Bill and Melinda Gates Foundation funded the acquisition of the data for this analysis by the IHME. The data were provided to the authors as volunteer collaborators with IHME.

## Details of the role of the study sponsors

While IHME GBD faculty and staff by virtue of Gates Foundation grants provided the raw data for this analysis, they did not vet the analysis or sponsor the manuscript.

## Statement of independence of researchers from funders

The researchers have received no funding. Gates Foundation funded IHME to collect and analyse the GBD data.

## Dissemination declaration

Dissemination of this manuscript to the participants of the more than 12,000 surveys is not possible individually, but the information will become in the public domain.

## Disclosures

We thank Scott Glenn and Brent Bell from IHME who supplied us with the GBD risk factor exposure data for the risk factors and for noncommunicable disease death data.

## Ethics

Studies were based solely on data from the IHME GBD database and do not need approval from any bioethics committee.

## Participant informed consents

Not applicable.

## Author access to data

As volunteer collaborators with the Institute of Health Metrics and Evaluation, we received about 1.4 gigabytes of raw data on noncommunicable disease deaths and 32 relevant risk factors.

## Protocol, submitted as a supplementary file

Not applicable.

## STROBE checklist

Submitted.

## Patient consent

Not applicable.

## Notes

### Competing Interest Statement

The authors have declared no competing interest.

### Clinical Trial

NA

