## Supplementary material for "Global Burden of Disease worldwide statistical analysis of risk factors impacting noncommunicable disease deaths compared with assessments by systematic literature reviews": Strobe checklist

**STROBE 2007 (v4) Statement—Checklist of items that should be included in reports of *cohort studies***

| **Section/Topic** | Item # | Recommendation | Reported on page # |
| --- | --- | --- | --- |
| **Title and abstract** | 1 | (*a*) Indicate the study’s design with a commonly used term in the title or the abstract | #1 |
|  |  | (*b*) Provide in the abstract an informative and balanced summary of what was done and what was found | #2-3 |
| Introduction | | |  |
| Background/rationale | 2 | Explain the scientific background and rationale for the investigation being reported | 6-7 |
| Objectives | 3 | State specific objectives, including any prespecified hypotheses | 7 |
| Methods | | |  |
| Study design | 4 | Present key elements of study design early in the paper | 8-10 |
| Setting | 5 | Describe the setting, locations, and relevant dates, including periods of recruitment, exposure, follow-up, and data collection | 8-10 |
| Participants | 6 | (*a*) Give the eligibility criteria, and the sources and methods of selection of participants. Describe methods of follow-up | 8-9 |
|  |  | (*b*) For matched studies, give matching criteria and number of exposed and unexposed | NA |
| Variables | 7 | Clearly define all outcomes, exposures, predictors, potential confounders, and effect modifiers. Give diagnostic criteria, if applicable | 10 and Supplementary Table 1 |
| Data sources/ measurement | 8 | For each variable of interest, give sources of data and details of methods of assessment (measurement). Describe comparability of assessment methods if there is more than one group | 10 and Supplementary Tables 2-3 |
| Bias | 9 | Describe any efforts to address potential sources of bias | 8 |
| Study size | 10 | Explain how the study size was arrived at | 9 |
| Quantitative variables | 11 | Explain how quantitative variables were handled in the analyses. If applicable, describe which groupings were chosen and why | 10 |
| Statistical methods | 12 | (*a*) Describe all statistical methods, including those used to control for confounding | 10-11 |
|  |  | (*b*) Describe any methods used to examine subgroups and interactions | 10 |
|  |  | (*c*) Explain how missing data were addressed | 8-9 |
|  |  | (*d*) If applicable, explain how loss to follow-up was addressed | NA |
|  |  | (*e*) Describe any sensitivity analyses | 10-11 |
| Results | | |  |
| Ecological dataↆ | 13 | (a) Report numbers of individuals at each stage of study—eg numbers potentially eligible, examined for eligibility, confirmed eligible, included in the study, completing follow-up, and analysed | 12-15 |
|  |  | (b) Give reasons for non-participation at each stage | NA |
|  |  | (c) Consider use of a flow diagram | NA |
| Descriptive data | 14 | (a) Give characteristics of study participants (eg demographic, clinical, social) and information on exposures and potential confounders | 12-15ↆ |
|  |  | (b) Indicate number of participants with missing data for each variable of interest | 8-9 |
|  |  | (c) Summarise follow-up time (eg, average and total amount) | 8 |
| Outcome data | 15 | Report numbers of outcome events or summary measures over time | 12-15, Tables 1-6 pages 24-32 |
| Main results | 16 | (*a*) Give unadjusted estimates and, if applicable, confounder-adjusted estimates and their precision (eg, 95% confidence interval). Make clear which confounders were adjusted for and why they were included | 12-15, Tables 1-6 pages 24-32 |
|  |  | (*b*) Report category boundaries when continuous variables were categorized | NA |
|  |  | (*c*) If relevant, consider translating estimates of relative risk into absolute risk for a meaningful time period | NA |
| Other analyses | 17 | Report other analyses done—e.g., analyses of subgroups and interactions, and sensitivity analyses | 14-15, Table 6 |
| Discussion |  |  |  |
| Key results | 18 | Summarize key results with reference to study objectives | 16-24 |
| **Limitations** |  |  | 25 |
| Interpretation | 20 | Give a cautious overall interpretation of results considering objectives, limitations, multiplicity of analyses, results from similar studies, and other relevant evidence | 25 |
| Generalisability | 21 | Discuss the generalisability (external validity) of the study results | 25 |
| Other information |  | Data availability statement | 38 |
| Funding | 22 | Give the source of funding and the role of the funders for the present study and, if applicable, for the original study on which the present article is based | 37 |

ↆGlobal Burden of Disease ecological data on worldwide male and female cohorts from IHME is the data source rather than individual participants
